# ASSOCIATIONS OF AUTISTIC TRAITS AND AUTISM WITH INCONTINENCE AND CONSTIPATION IN A UK BIRTH COHORT

**DOI:** 10.1101/2024.10.02.24314806

**Authors:** Prince Gyamenah, Kimberley Burrows, Dheeraj Rai, Carol Joinson

**Affiliations:** Epidemiology, Population Health Sciences, Bristol Medical School, University of Bristol, United Kingdom; Senior Research Associate in Epidemiology, Bristol Medical School, University of Bristol, United Kingdom; Professor of Neurodevelopmental Psychiatry, Population Health Sciences, Bristol Medical School, University of Bristol, United Kingdom; Professor of Developmental Psychology, Bristol Medical School, University of Bristol, United Kingdom

**Keywords:** Autistic traits/autism, incontinence/constipation, sociability, repetitive behaviours, social-communication, speech coherence, and ALSPAC

## Abstract

**Background:** There is evidence that children with autism/autistic traits have higher risks of incontinence and constipation, but no studies have examined this in a large community-based cohort.

**Aim/Research question:** are autistic traits and diagnosed autism prospectively associated with increased odds of incontinence and constipation in children and adolescents?

**Methods:** This was a population-based cohort study based on data from the Avon Longitudinal Study of Parents and Children (n=4233-4490 at age 9 years; n=3403-3697 at age 14). We used multivariable logistic regression to examine associations of parent-reported autistic traits (sociability, repetitive behaviours, social-communication, coherence) (at ages 3-9 years) and autism with incontinence (bedwetting, daytime-wetting, soiling) and constipation (parent-reported at age 9, self-reported at age 14). We adjusted for parity, maternal age at delivery, child’s sex and developmental level, maternal depression, and anxiety (antenatal and postnatal), and indicators of family socioeconomic status.

**Results:** Social-communication and speech coherence difficulties showed the strongest associations with incontinence, e.g. adjusted odds ratio (OR) and 95% confidence interval (CI) for the association between social-communication difficulties and daytime-wetting was 2.21 (1.47-3.32) and for coherence was 2.34 (1.60-3.43). The odds of soiling were also higher in children with social-communication (OR:1.88, 95%CI:1.28-2.75) and coherence difficulties (OR:2.04, 95%CI:1.43-2.93). Diagnosed autism was only associated with an increase in the odds of daytime-wetting (OR:3.18, 95%CI:1.44-7.02). At 14 years, there was less evidence of associations between autistic traits and incontinence but there was evidence of associations between autistic traits and constipation: social-communication (OR:1.68, 95%CI:1.13-2.49), coherence difficulties (OR:1.64, 95%CI:1.11-2.41). 5

**Conclusions:** Early assessment and treatment of incontinence/constipation should be considered for children with autistic traits.

## INTRODUCTION

Autistic traits that are commonly seen in individuals with Autism Spectrum Disorder (henceforth autism) include difficulties with social interaction and communication, repetitive behaviours, and restricted interests (Nazeer & Ghaziuddin, 2012). Individuals in the community who do not meet the diagnostic criteria for autism can still exhibit autistic traits since these traits are continuously distributed in the population (Marinopoulou et al., 2023). The prevalence of autism has increased over the past few decades with an estimated prevalence of 1 to 2% in the general population (Niemczyk et al., 2018).

There is evidence that continence problems are more common among children with autism/autistic traits than in the general population. For example, the prevalence of incontinence among children with autism/autistic traits (mean age 11.3 years) compared with controls (mean age 10.7 years) was estimated to be 30% versus 0% for bedwetting, 45% versus 4.7% for daytime wetting, and 42.5% versus 7.5% for children with delayed bowel control (Gubbiotti et al., 2019; von Gontard et al., 2015). Factors that are associated with incontinence in children with autism/autistic traits include delayed development of bladder control, cognitive deficits, and behavioural challenges such as resistance to toilet training (Dalrymple & Ruble, 1992; Niemczyk et al., 2018; Raturi et al., 2021; Tu et al., 2017).

Constipation is also commonly observed in children with autism/autistic traits. A recent review revealed that the rates of constipation among children with autism range from 4.3 to 45% with a median of 22% (Mulay & Karthik, 2022). The prevalence of childhood constipation in the general population has been reported to range from 0.7% to 29% (Rajindrajith et al., 2016). The aetiology of constipation in children with autism/autistic traits could be multifactorial, with factors including genetic predisposition, low socioeconomic status, inadequate daily fibre intake, and insufficient fluid intake. Another common factor for constipation in children with autism/autistic traits could be stool withholding arising from a previous painful or distressing bowel movement (McElhanon et al., 2014; Mugie et al., 2011).

Previous studies have found associations between autism/autistic traits and incontinence/constipation, but some studies are limited by small sample sizes and inadequate adjustments for potential confounders (Gubbiotti et al., 2019; Harris et al., 2021; Niemczyk et al., 2019; Peters et al., 2014b). A cross-sectional study of children aged 2-17 years with autism (selected from the Autism Treatment Network in the United States), found that rigid or compulsive behaviours are associated with severe constipation and co-occurring diarrhoea/underwear staining, after adjusting for covariates including non-verbal IQ, anxiety, depression, and Attention-Deficit-Hyperactivity-Disorder (ADHD) (Peters et al., 2014a). A case-control study of 51 children with autism (43 boys and 8 girls, mean age = 9.7 years) and 53 matched controls (43 boys and 10 girls, mean age = 10.2 years) found that children with autism have a higher prevalence of bedwetting and daytime wetting compared with typically developing children (Niemczyk et al., 2019). The study was conducted in a specialized outpatient clinic for autism, which may limit the generalizability of the findings to other settings. Population-based prospective studies have also reported associations between autism/autistic traits and constipation. These studies, however, did not adjust for potential confounders including child development level, social class, and maternal psychopathology (Gubbiotti et al., 2019; Harris et al., 2021). (Supplementary Table 1 includes further details of the studies reviewed here).

To our knowledge, no prospective cohort study has examined if specific autistic traits are differentially associated with incontinence/constipation in children and adolescents. This is important because certain autistic traits could identify children at greater risk of chronic incontinence/constipation. Parents and carers of children with autistic traits could be provided with guidance and advice on toileting, diet and fluid intake to reduce the risk of continence problems and constipation. The aim of this study is to examine prospective associations of autistic traits (social-communication, coherence, repetitive behaviours, and sociability) and diagnosed ASD with incontinence (bedwetting, daytime-wetting, and soiling) and constipation in children (9 years) and adolescents (14 years).

## METHODOLOGY

### STUDY DESIGN, SETTING, AND PARTICIPANTS

This was a large population-based prospective cohort study using participants from the Avon Longitudinal Study of Parents and Children (ALSPAC) cohort in the United Kingdom. ALSPAC is a longitudinal birth cohort of children born in the former county of Avon, in the UK. Pregnant women resident in Avon with expected dates of delivery between 1st April 1991 and 31^st^ December 1992 were invited to take part in the study. The initial number of pregnancies enrolled was 14541. Of the initial pregnancies, there was a total of 14676 fetuses, resulting in 14062 live births and 13988 children who were alive at 1 year of age. When the oldest children were approximately 7 years of age, an attempt was made to bolster the initial sample with eligible cases who had failed to join the study originally. The total sample size for analyses using any data collected after the age of seven is therefore 15447 pregnancies, resulting in 15658 fetuses. Of these, 14901 children were alive at 1 year of age (Boyd et al., 2013; Fraser et al., 2013).

The ALSPAC cohort has comprehensive data on both parents and children, which was collected prospectively across several occasions during pregnancy and throughout the developmental period of childhood. The sources of data include self-reported questionnaires, clinical evaluations, medical history, and educational records. At age 18, study children were sent ‘fair processing’ materials describing ALSPAC’s intended use of their health and administrative records and were given clear means to consent or object via a written form. Data were not extracted for participants who objected, or who were not sent fair processing materials (Rai et al., 2018). The study website contains details of all the data that is available through a fully searchable data dictionary and variable search tool at http://www.bristol.ac.uk/alspac/researchers/our-data/.

### ETHICAL APPROVAL AND INFORMED CONSENT

Ethical approval for the study was obtained from the ALSPAC Ethics and Law Committee and the Local Research Ethics Committees. Precise details on the ethics committee/institutional review board(s) are provided at this link: http://www.bristol.ac.uk/alspac/researchers/research-ethics/. Informed consent for the use of data collected via questionnaires and clinics was obtained from participants following the recommendations of the ALSPAC Ethics and Law Committee at the time.

### ASSESSMENT OF AUTISTIC TRAITS

When the study children were aged 11 years, ALSPAC had collected 93 measures related to autistic traits. Four individual measures were previously found to be the strongest predictors of ASD. These were the sociability subscale of the Emotionality Activity and Sociability temperament measure at 3 years (supplementary text 1), an ALSPAC-defined repetitive behaviour scale at 5 years (supplementary text 2), the Social Communication Disorders Checklist (SCDC) at 7 years (supplementary text 3), and the coherence subscale of the Children’s Communication Checklist at 9 years (supplementary text 4). We also used a factor-mean-score(FMS), which is a composite measure of all of the autistic traits (supplementary text 5). We used this to allow us to examine if a broad autism phenotype is associated with an increased risk of incontinence and constipation. We dichotomized the autistic traits and the FMS into cases and non-cases. Consistent with previous ALSPAC-based research, we dichotomized each ASD trait into a high-risk group (for ASD) which was as close as possible to 10% of the population. This threshold has been used in previous studies and shown to be predictive of ASD. Dichotomizing autistic traits helps to identify high-risk groups whilst providing sufficient statistical power for analysis (Guyatt et al., 2015; Rai et al., 2018; Steer et al., 2010).

### ASCERTAINMENT OF ASD CASES

Diagnosis of autism is currently made through assessment by a multidisciplinary team and involves history-taking from parents/carers and observation of the child. A multi-source approach was used to accurately identify children in the ALSPAC cohort with an autism diagnosis. This approach involved an examination of clinical records for all children who underwent a multidisciplinary assessment for a developmental disorder, which was validated against the International Statistical Classification of Diseases, 10th Revision (ICD-10) criteria by a consultant paediatrician. Additionally, special education support records for autism were reviewed, and parental reports of autism or Asperger syndrome diagnosis were considered (Rai et al., 2018; Williams et al., 2008). Cases were individuals who had confirmed diagnosis of autism while non-cases were those who did not have an autism diagnosis.

### ASSESSMENT OF INCONTINENCE/CONSTIPATION

The International Children’s Continence Society and the Rome Criteria provide globally accepted guidelines for the evaluation of paediatric incontinence and constipation. However, consistent with other community-based studies, we did not use diagnostic criteria for incontinence and constipation because the number of participants who experienced severe incontinence and constipation is small compared with clinical samples. Restricting the analysis to those children who met clinical diagnostic criteria would have resulted in low statistical power and a lack of precision in our estimates. We therefore examined the presence versus absence of any incontinence and constipation. It is important to note that we found robust associations between autistic traits and incontinence/constipation even when examining cases that did not meet the criteria for clinical diagnosis (Austin et al., 2016; Chang et al., 2017; Hyams et al., 2016).

In our study, incontinence and constipation were reported in questionnaires by mothers when their study children were aged 9 years and self-reported at age 14 years. When study children were aged 9 years, their parents were asked “How often does your child wet the bed at night; wet his/herself during the day; dirty his/her pants during the day?” Responses included “1-Never”, “2-Occasional accidents less than once a week”, “3-About once a week”, “4-Two to five times a week”, “5-Nearly every day” and “6-More than once a day”. We categorised “1-Never” as non-cases and responses 2 to 6 as cases. For constipation, parents were asked, “Has he/she had any constipation in the past 12 months?” Responses included “1-Yes, saw a doctor”, “2-Yes, did not see a doctor”, and “3-No, did not have”. We categorised responses 1 and 2 as cases and response 3 as non-cases.

Similarly, when the study children were aged 14 years, they were asked “How often do you wet the bed at night; wet yourself during the day; dirty your pants during the day?” Responses included “1-Never”, “2-Occasional accidents less than once a week”, “3-About once a week”, “4-Two to five times a week”, “5-Nearly every day” and “6-More than once a day”. We categorised “1-Never” as non-cases and responses 2 to 6 as cases. For constipation, the children were asked, “Have you had any constipation in the past 12 months?” Responses included “1-Yes, saw a doctor”, “2-Yes, did not see a doctor”, and “3-No, did not have”. We categorised responses 1 and 2 as cases and response 3 as non-cases.

### CONFOUNDERS/COVARIATES

We adjusted for potential confounders that were chosen because of existing evidence that they are associated with autism/autistic traits and incontinence/constipation. Confounders included parity, maternal age at delivery, child’s sex, child’s developmental level at 18 months, maternal depression (antenatal, measured at 18 weeks and 32 weeks of gestation and postnatal, measured at 8 weeks and 8 months after delivery), maternal anxiety (antenatal, measured at 18 weeks of gestation and postnatal, measured at 8 weeks after delivery), family social class, home ownership status, highest educational attainment, and financial difficulty (Fernandes, 2015; Niemczyk et al., 2018; Peters et al., 2014a; Rai et al., 2018). (Details of the confounders are found in supplementary table 2).

### STATISTICAL ANALYSIS

We used multivariable logistic regression models and adjusted for the confounders. We also performed descriptive statistics and assessed the differences in proportions using Chi^2^. All analyses were conducted in Stata 17 (StataCorp. 2021).

## RESULTS

Participants with complete data for each exposure variable (autistic traits/autism diagnosis) together with outcome variables (incontinence/constipation) and confounders ranged from 4233 to 4490 at age 9 years, and 3403 to 3697 at age 14 years (see the study flow diagram in figure 1). The sample (n=4233) at age 9 years with complete data (for at least one autistic trait, incontinence/constipation, and confounders), had a lower proportion of manual social class, lowest level of maternal education, and parity of two or more compared with the other samples as shown in supplementary table 3, but the proportion of males was similar across the samples. A similar distribution of characteristics was found at age 14 years for those with complete data compared with the other samples (supplementary table 4). At age 9 years, 84% of children who had social-communication difficulties were in the non-manual social class (supplementary table 5). Similarly, at age 14 years, 86% of children who had social-communication difficulties were in the non-manual social class (supplementary table 6).

**Fig. 1.**
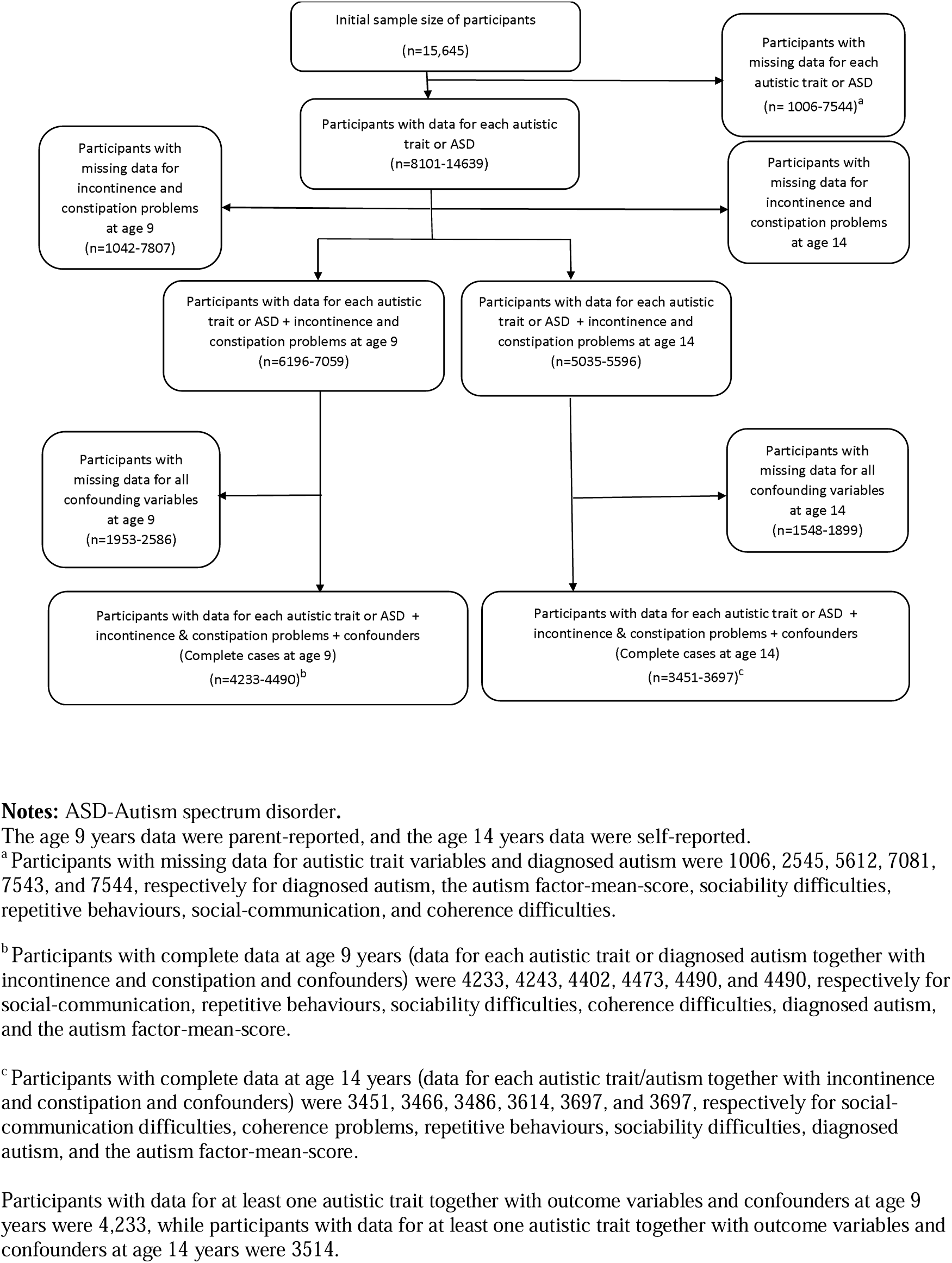
Flow chart showing participants included in the study.

Social-communication and coherence showed the strongest evidence of associations with daytime wetting and soiling at age 9 years in the adjusted models (table 1). Difficulties with social-communication and coherence were associated with an increase in the odds of daytime wetting (odds ratio (OR):2.21, 95% confidence interval (CI):1.47-3.32) and OR:2.34, CI:1.60-3.43, respectively) and with an increase in the odds of soiling (OR:1.88, CI:1.28-2.75 and OR:2.04, CI:1.43-2.93, respectively). The adjusted associations of repetitive behaviour and sociability with daytime wetting and soiling crossed the null value. We found evidence of associations between the autism factor-mean-scores and daytime wetting (OR:1.76, CI:1.14-2.71), and soiling (OR:2.18, CI:1.51-3.15), after adjusting for confounders. Diagnosed autism was only associated with an increase in the odds of daytime wetting (OR:3.18 CI:1.44-7.02). There was weaker evidence of associations between autistic traits/autism diagnosis and bedwetting: associations were only observed for coherence difficulties (OR:1.37, CI:1.01-1.86) and the factor-mean-scores (OR:1.33, CI:1.01-1.81). There was little evidence that the autistic traits/autism diagnosis were associated with increased odds of constipation at age 9 years, except for an association with the autism factor-mean-score (OR:1.57, CI:1.11-2.20)

**Table 1.** The associations between autistic traits/diagnosed autism and incontinence/constipation problems in children at age 9 years.

| Variables | Sample size (N) | Unadjusted estimates |  |  | Adjusted estimates* |  |  |
| --- | --- | --- | --- | --- | --- | --- | --- |
|  |  | OR | 95% CI | P-value | OR | 95% CI | P-value |
| Bedwetting |  |  |  |  |  |  |  |
| Social communication | 4233 | 1.73 | 1.28- 2.34 | <0.001 | 1.32 | 0.96- 1.81 | 0.08 |
| Coherence | 4473 | 1.68 | 1.26-2.25 | <0.001 | 1.37 | 1.01-1.86 | 0.04 |
| Repetitive behaviour | 4243 | 1.40 | 0.97-2.00 | 0.07 | 1.14 | 0.79-1.65 | 0.49 |
| Sociability | 4402 | 1.22 | 0.91-1.64 | 0.18 | 1.09 | 0.81-1.49 | 0.54 |
| Factor-mean-score<sup>a</sup> | 4490 | 1.89 | 1.41-2.53 | <0.001 | 1.33 | 1.01-1.82 | 0.05 |
| Diagnosed autism | 4490 | 2.23 | 1.14-4.33 | 0.02 | 1.70 | 0.86-3.37 | 0.13 |
| Daytime wetting |  |  |  |  |  |  |  |
| Social communication | 4233 | 2.41 | 1.64- 3.54 | <0.001 | 2.21 | 1.47-3.32 | <0.001 |
| Coherence | 4473 | 2.39 | 1.67-3.43 | <0.001 | 2.34 | 1.60-3.43 | <0.001 |
| Repetitive behaviour | 4243 | 1.61 | 1.00-2.60 | 0.05 | 1.47 | 0.90-2.40 | 0.13 |
| Sociability | 4402 | 1.03 | 0.67-1.60 | 0.89 | 1.10 | 0.71-1.72 | 0.66 |
| Factor-mean-score<sup>a</sup> | 4490 | 1.94 | 1.31- 2.87 | 0.001 | 1.76 | 1.14-2.71 | 0.01 |
| Diagnosed autism | 4490 | 3.33 | 1.56-7.13 | 0.002 | 3.18 | 1.44-7.02 | 0.004 |
| Soiling |  |  |  |  |  |  |  |
| Social communication | 4233 | 2.50 | 1.74-3.59 | <0.001 | 1.88 | 1.28-2.75 | 0.001 |
| Coherence | 4473 | 2.67 | 1.90-3.75 | <0.001 | 2.04 | 1.43-2.93 | <0.001 |
| Repetitive behaviour | 4243 | 1.58 | 1.00-2.51 | 0.05 | 1.28 | 0.80-2.05 | 0.31 |
| Sociability | 4402 | 1.36 | 0.93-1.99 | 0.11 | 1.29 | 0.87-1.90 | 0.20 |
| Factor-mean-score<sup>a</sup> | 4490 | 3.24 | 2.33-4.50 | <0.001 | 2.18 | 1.51-3.15 | <0.001 |
| Diagnosed autism | 4490 | 3.01 | 1.41-6.43 | 0.004 | 2.10 | 0.96-4.62 | 0.06 |
| Constipation |  |  |  |  |  |  |  |
| Social communication | 4233 | 1.29 | 0.92-1.80 | <0.14 | 1.20 | 0.85-1.69 | 0.30 |
| Coherence | 4473 | 1.16 | 0.83-1.61 | 0.38 | 1.15 | 0.81-1.62 | 0.43 |
| Repetitive behaviour | 4243 | 1.07 | 0.72-1.59 | 0.74 | 0.98 | 0.66-1.47 | 0.94 |
| Sociability | 4402 | 0.86 | 0.61-1.20 | 0.37 | 0.91 | 0.65-1.27 | 0.58 |
| Factor-mean-score<sup>a</sup> | 4490 | 1.56 | 1.15-2.13 | 0.01 | 1.57 | 1.11-2.20 | 0.01 |
| Diagnosed autism | 4490 | 0.92 | 0.36-2.31 | 0.86 | 0.92 | 0.36-2.34 | 0.86 |

\*
The confounders adjusted for include parity, maternal age at delivery, child's sex, child's developmental level at 18 months, maternal depression (antenatal and postnatal), maternal anxiety (antenatal and postnatal), family social class, home ownership status, highest educational attainment, and financial difficulty.
<sup>a</sup>The factor-mean-score is a broad autism phenotype, and it is a composite measure of all of the autistic traits (Steer et al., 2010).

In contrast to the results at age 9 years, there was less evidence of associations between autistic traits and incontinence at age 14 years in the adjusted models (table 2). The autism factor-mean-score was associated with increased odds of bedwetting (OR:2.59, CI:1.40-4.81), daytime-wetting (OR:2.26, CI:1.20-4.26), and soiling (OR:2.46, CI:1.52-3.97). Coherence difficulties and diagnosed autism were associated with an increase in the odds of soiling at age 14 years (OR:1.77, CI:1.10-2.85 and OR:3.81, CI:1.40-10.34, respectively). In comparison to the results at age 9, there was evidence of associations between the autistic traits and constipation: social-communication (OR:1.68, CI:1.13-2.49), coherence difficulties (OR:1.64, CI:1.11-2.41), repetitive behaviours (OR:1.88, CI:1.24-2.87), autism factor-mean-score (OR:1.53, CI:1.01-2.32).

**Table 2.** The associations between autistic traits/diagnosed autism and incontinence/constipation problems in children at age 14 Years.

| Variables | Sample size (N) | Unadjusted estimates |  |  | Adjusted estimates<sup>*</sup> |  |  |
| --- | --- | --- | --- | --- | --- | --- | --- |
|  |  | OR | 95% CI | P-value | OR | 95% CI | P-value |
| Bedwetting |  |  |  |  |  |  |  |
| Social communication | 3451 | 1.24 | 0.59-2.60 | 0.57 | 1.13 | 0.53-2.40 | 0.75 |
| Coherence | 3466 | 1.67 | 0.87-3.19 | 0.12 | 1.57 | 0.80-3.08 | 0.19 |
| Repetitive behaviour | 3486 | 0.37 | 0.09-1.52 | 0.17 | 0.35 | 0.08-1.43 | 0.14 |
| Sociability | 3614 | 1.02 | 0.51-2.06 | 0.95 | 0.95 | 0.47-1.91 | 0.88 |
| Factor-mean-score<sup>a</sup> | 3697 | 2.85 | 1.63-4.98 | <0.001 | 2.59 | 1.40-4.81 | 0.002 |
| Diagnosed autism | 3697 | 1.36 | 0.18-10.06 | 0.76 | 1.18 | 0.16-8.87 | 0.87 |
| Daytime wetting |  |  |  |  |  |  |  |
| Social communication | 3451 | 1.17 | 0.58-2.35 | 0.65 | 1.07 | 0.52-2.23 | 0.85 |
| Coherence | 3466 | 1.41 | 0.76-2.60 | 0.28 | 1.43 | 0.74-2.74 | 0.30 |
| Repetitive behaviour | 3486 | 0.80 | 0.32-1.98 | 0.62 | 0.86 | 0.34-2.17 | 0.75 |
| Sociability | 3614 | 1.02 | 0.54-1.92 | 0.95 | 1.07 | 0.56-2.03 | 0.85 |
| Factor-mean-score<sup>a</sup> | 3697 | 2.01 | 1.15-3.52 | 0.14 | 2.26 | 1.20-4.26 | 0.01 |
| Diagnosed autism | 3697 | 1.08 | 0.15-8.00 | 0.94 | 1.24 | 0.16-9.56 | 0.83 |
| Soiling |  |  |  |  |  |  |  |
| Social communication | 3451 | 1.53 | 0.92-2.55 | 0.10 | 1.41 | 0.83-2.39 | 0.20 |
| Coherence | 3466 | 1.97 | 1.25-3.09 | 0.003 | 1.77 | 1.10-2.85 | 0.02 |
| Repetitive behaviour | 3486 | 1.37 | 0.76-2.46 | 0.29 | 1.38 | 0.76-2.50 | 0.29 |
| Sociability | 3614 | 1.55 | 0.99-2.43 | 0.06 | 1.56 | 0.99-2.47 | 0.05 |
| Factor-mean-score<sup>a</sup> | 3697 | 2.60 | 1.70-3.97 | <0.001 | 2.46 | 1.52-3.97 | <0.001 |
| Diagnosed autism | 3697 | 4.03 | 1.53-10.60 | 0.005 | 3.81 | 1.40-10.34 | 0.01 |
| Constipation |  |  |  |  |  |  |  |
| Social communication | 3451 | 1.90 | 1.30-2.80 | 0.001 | 1.68 | 1.13-2.49 | 0.01 |
| Coherence | 3466 | 1.87 | 1.29-2.71 | 0.001 | 1.64 | 1.11-2.41 | 0.01 |
| Repetitive behaviour | 3486 | 2.15 | 1.43-3.23 | <0.001 | 1.88 | 1.24-2.87 | 0.003 |
| Sociability | 3614 | 1.35 | 0.93-1.96 | 0.11 | 1.35 | 0.93-1.98 | 0.12 |
| Factor-mean-score<sup>a</sup> | 3697 | 1.83 | 1.25-2.68 | 0.002 | 1.53 | 1.01-2.32 | 0.05 |
| Diagnosed autism | 3697 | 2.37 | 0.91-6.22 | 0.08 | 2.27 | 0.85-6.03 | 0.10 |

<sup>\*</sup>
The confounders adjusted for include parity, maternal age at delivery, child's sex, child's developmental level at 18 months, maternal depression (antenatal and postnatal), maternal anxiety (antenatal and postnatal), family social class, home ownership status, highest educational attainment, and financial difficulty.
<sup>a</sup>The factor-mean-score is a broad autism phenotype, and it is a composite measure of all of the autistic traits (Steer et al., 2010).

## DISCUSSION

This study is the first to assess the association of different autistic traits and autism with incontinence/constipation in a large prospective cohort study. We found evidence that difficulties with social-communication and coherence were associated with increased odds of daytime wetting and soiling at age 9 years. We also found that the autism factor-mean-score (indicative of a broad autism phenotype) was associated with daytime wetting and soiling and diagnosed autism was associated with daytime wetting. There was weaker evidence of associations between the autistic traits and bedwetting at age 9, and little evidence of associations with constipation at age 9, except for the autism factor-mean-score. At age 14 years, the associations between the autism factor-mean-score and incontinence/constipation persisted and associations were also found between coherence difficulties, autism diagnosis and soiling. We found less evidence of associations between autistic traits/diagnosed autism and incontinence at age 14, but autistic traits were associated with constipation at age 14 years.

Our findings are consistent with previous studies which have found associations between autism/autistic traits and incontinence/constipation, but these studies were either restricted to specialised settings such as paediatric clinics, had small sample sizes, or used a cross-sectional design (Cuffman & Burkhart, 2021; Gubbiotti et al., 2019; Niemczyk et al., 2019; Peters et al., 2014a). Strengths of our study include the prospective birth cohort design and the availability of data on diagnosed autism in addition to different autistic traits. The autistic traits we examined have previously been found to have good predictive validity for autism and enabled us to examine if there are differential associations with incontinence/constipation (Rai et al., 2018). Other strengths include the availability of data on both parent-reported (in late childhood) and self-reported (in adolescence) incontinence/constipation, and a range of confounders. This large community-based cohort includes a majority of the children in the population who do not meet the diagnostic criteria for autism. This is important because autistic traits are believed to be on a continuum which means that our findings generalise beyond children with a clinical diagnosis of autism (Marinopoulou et al., 2023).

Although we adjusted for a range of potential confounders, the possibility of residual and unmeasured confounding cannot be ruled out, which limits our ability to infer causality. This study, like other cohort studies, suffered from missing data and attrition, which could potentially cause selection bias since the complete case sample was more socially advantaged than the original cohort. Our estimates, however, should be unbiased provided there are no systematic differences in the rates of the outcomes considered after conditioning on the exposure and confounders included in the model. Since we accounted for a number of confounders including known predictors of missing data in ALSPAC, and childhood incontinence in ALSPAC has been shown to be only weakly socially patterned, we believe this assumption is tenable in this current study (Butler & Heron, 2008; Hughes et al., 2019).

Compared with the autistic traits, our study had lower statistical power to examine associations between diagnosed autism and incontinence/constipation due to the small number of autism cases in this community-based sample (see supplementary tables 5 and 6). This is reflected in the imprecise estimates with wide confidence intervals.

Another limitation of this study is the use of self-reported questionnaires for autistic traits and incontinence/constipation. However, differential misclassification is unlikely because this is a prospective study, and the outcomes (incontinence/constipation) were not known when parents provided data on the autistic traits. The exception is the coherence subscale of the Children’s Communication Checklist, which was completed at 9 years, which might explain why associations of this autistic trait with incontinence at age 9 were stronger. It is notable, however, that coherence difficulties at age 9 were prospectively associated with soiling and constipation at age 14.

It is possible that the stronger associations of coherence difficulties and social-communication with incontinence may be attributed to the concept of fractionation of component features of autism. This concept suggests that social and non-social aspects of autism may have distinct causes, and hence may present with different symptoms. The timing of assessments of autistic traits (sociability and repetitive behaviours) could also have affected the strength of the associations. Compared with social-communication and coherence difficulties (which were reported at ages 7 and 9 respectively), the sociability and repetitive behaviour scales were completed when the children were younger (at ages 3 and 5 years), hence certain features might not have been fully developed or expressed. The stronger associations between autistic traits and constipation at age 14 years than at age 9 years could be due to self-reporting of constipation at age 14 years by the study children, compared with the use of parent reports at age 9 which might have missed constipation (Happé & Ronald, 2008; Joinson et al., 2019).

Developmental delays in some children with autism may lead to difficulties with toilet training which could explain the associations between autistic traits/autism and incontinence (Francis et al., 2017). Stool withholding, which can arise from a previous painful or distressing bowel movement is often common in children with autism and this is associated with constipation (Raturi et al., 2021). Children with autistic traits often also have neurological and sensory processing difficulties that affect how they perceive and respond to bodily sensations, including those related to bladder and bowel functions (McElhanon et al., 2014; Woodward, 1996).

Previous studies have indicated that children with autistic traits often have restricted diets or preferences for starches, snack foods, and processed foods and avoid certain textures or types of foods including fruits, vegetables, and proteins (Harris et al., 2021; McElhanon et al., 2014). It is well established that diets deficient in fibre increase the risk of constipation (Cummings, 1984; Mugie et al., 2011). Studies have also found evidence that children with autistic traits may have altered microbiomes, which might be a consequence of their restricted diet (Dan et al., 2020; Son et al., 2015). Alterations of intestinal microbiota may contribute to constipation and constipation-related symptoms (Zhang et al., 2021).

Genetic factors might also explain the associations between autistic traits/autism and incontinence. Studies suggest that autistic traits may result from many different patterns of genetic causality including single genes, combinations of several genes, and large groups of genes (Wei et al., 2021). Consequently, there could be shared genes for autistic traits and incontinence/constipation, which could be a common underlying cause.

This population-based study provides new evidence that children with autistic traits/autism are more likely to experience subsequent problems with incontinence/constipation. Social-communication and coherence difficulties were more strongly associated with incontinence than the other autistic traits. Incontinence and constipation can significantly impact a person’s quality of life, leading to embarrassment, social isolation, emotional distress, a loss of confidence, and also have an impact on mental health (Flaherty, 2019; Grzeda et al., 2017). Early assessment and treatment for incontinence and constipation should be considered for children with autistic traits to reduce the risk of incontinence/constipation becoming chronic.

## Data Availability

The study website contains details of all the data that is available through a fully searchable data dictionary and variable search tool at http://www.bristol.ac.uk/alspac/researchers/our-data/.

## Acknowledgements

We are extremely grateful to all the families who took part in this study, the midwives for their help in recruiting them, and the whole ALSPAC team, which includes interviewers, computer and laboratory technicians, clerical workers, research scientists, volunteers, managers, receptionists, and nurses.

## SUPPLEMENTARY MATERIALS

### SUPPLEMENTARY TEXT 1

The sociability subscale of the Emotionality Activity and Sociability (EAS) temperament measure is used to assess an individual’s preference for being with others rather than being alone. It measures the extent to which an individual seeks social interaction, shares activities, and desires attention from others (Bould et al., 2013). The sociability subscale is one of the four subscales of the EAS measure, which also includes emotionality, activity, and impulsivity. The EAS measure is widely used to assess temperament in children and has been found to have good stability over time (Bould et al., 2013). The items are evaluated using a five-point Likert scale, which ranges from one (not typical) to five (very typical). The summation of the items within the scale is divided by five to derive the corresponding score. The items in the sociability subscale include.

1: Likes to be with people.

2: Prefers playing with others rather than alone.

3: Finds people more stimulating than anything else.

4: Is something of a loner.

5: When alone, the child feels isolated.

### SUPPLEMENTARY TEXT 2

The Repetitive Behaviour Scale is a tool used to assess and describe repetitive and restricted behaviours in various psychiatric syndromes, including autism spectrum disorders (Bourreau et al., 2009). It provides a standardized and accurate description of these behaviours, allowing for a better understanding of their clinical dimension (Bourreau et al., 2009). The scale has been found to have good psychometric qualities, including good interrater reliability, internal consistency, and content validity (Bourreau et al., 2009). The scale has potential clinical applications and can be used in research, treatment, and clinical practice. In the ALSPAC, this scale was developed from the answers to four questions in the questionnaire sent to the mother at 69 months and these were as follows.

a. How often does he/she repeatedly rock his head or body for no reason?
b. How often does he/she have a tic or twitch?
c. How often does he/she have other unusual behaviour?
d. How often does he/she stumble or get stuck on words, or repeat them many times? (example, I I I I want a sweet)? The responses to each question were coded as: “often/always =3”, “sometimes =2”, and “never =1”.

### SUPPLEMENTARY TEXT 3

The Social and Communication Disorders Checklist, also known as SCDC, is a twelve-item checklist that was designed to allow for quick and efficient evaluation of autistic traits. Within this set of questions, nine are directed towards measuring abnormalities in the aspects of the autistic triad that pertain to reciprocal social interaction and communication skills (Skuse et al., 2005). Each item on the scale is evaluated based on its occurrence within the past six months and whether the associated statements are “not true”, “quite or sometimes true” or “very or often true.” The assigned scores are “0”, “1”, or “2”, which culminate in a maximum score of twenty-four (Skuse et al., 2005). The SCDC has strong internal consistency and test-retest stability (Solmi et al., 2021). Items in the SCDC scale are as follows;

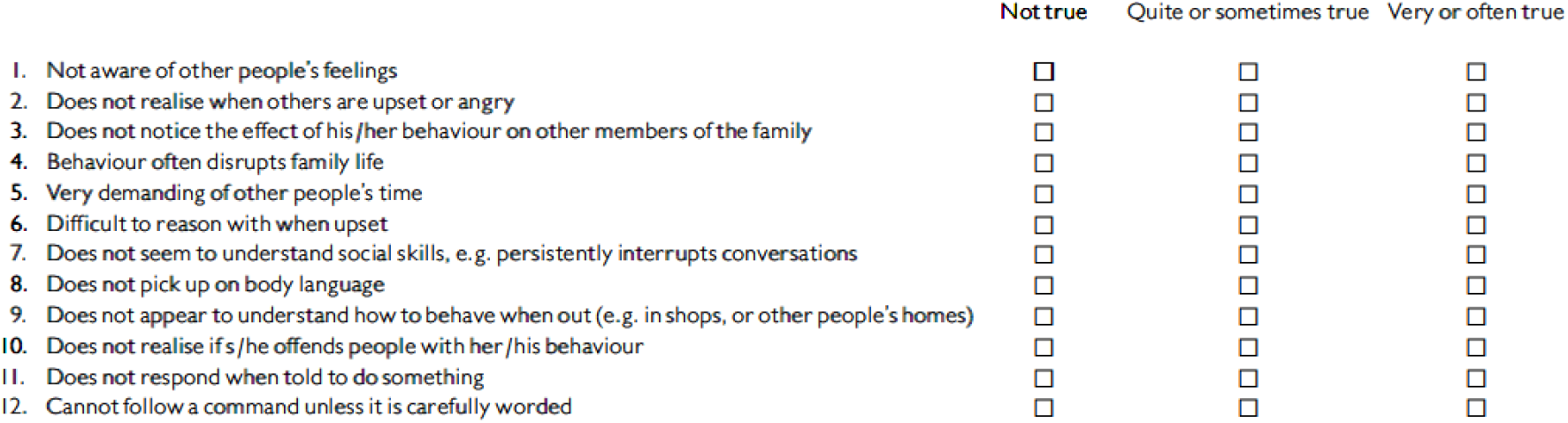

### SUPPLEMENTARY TEXT 4

The children communication checklist (CCC) is a standardized questionnaire comprising seventy items, which has been specifically developed to evaluate the communicative abilities of children (Lane et al., 2019). This instrument is highly effective in detecting significant communicative difficulties among children (Lane et al., 2019). The CCC-2 questionnaire consists of ten subscales, each of which assesses a different aspect of communication, including (A) speech, (B) syntax, (C) semantics, (D) coherence, (E) inappropriate initiation, (F) stereotyped language, (G) use of context, (H) nonverbal communication, (I) social relations, and (J) interests. Each of these subscales is comprised of seven items, five of which are related to communicative difficulties, and two of which are related to communicative strengths (Lane et al., 2019). Analyses of traits predictive of autism in ALSPAC showed that the coherence subscale of the CCC performed better than the other CCC scales (Golding et al., 2017), and therefore that was used in our study. The coherence scale is made up of eight items which are as follows.

1. It is sometimes hard to make sense of what he is saying because it seems illogical or disconnected.
2. Conversation with him can be enjoyable and interesting.
3. Can give an easy-to-follow account of a past event such as a birthday party or holiday.
4. Can talk clearly about what he plans to do in the future (example, tomorrow, or next week)
5. Would have difficulty in explaining to a younger child how to play a simple game such as ‘‘snap’’.
6. Has difficulty in telling a story, or describing what he has done, in an orderly sequence of events.
7. Uses terms like ‘‘he’’ or ‘‘it’’ without making it clear what he is talking about.
8. Doesn’t seem to realise the need to explain what he is talking about to someone who doesn’t share his experiences; for instance, might talk about ‘‘Johnny’’ without explaining who he is.

The items included in the questionnaire are evaluated using a Likert scale, which provides an assessment of the frequency of communicative problems and strengths. The scale ranges from zero, representing less than once a week or never, to three, representing several times a day or always (Lane et al., 2019).

### SUPPLEMENTARY TEXT 5

The factor-mean-score as a composite measure of autistic traits was derived using principal components analysis (PCA) (Steer et al., 2010). Ninety-three traits that potentially indexed the emergence and presence of autism were selected for factor analysis (Steer et al., 2010). The parental questionnaire, as well as observational and standardized tests, were used to select measures that were included in the factor analysis (Bolton et al., 2012). These measures were collected at various time points in development. The factor analysis identified seven principal factors which related to verbal ability, language acquisition, social understanding, semantic-pragmatic skills, repetitive stereotyped behaviour, articulation, and social inhibition (Bolton et al., 2012). A single common autistic trait score was derived by taking the mean score of these seven factors and low scores on this derived variable reflected more autistic traits (Bolton et al., 2012).

---

**Supplementary table 1.**
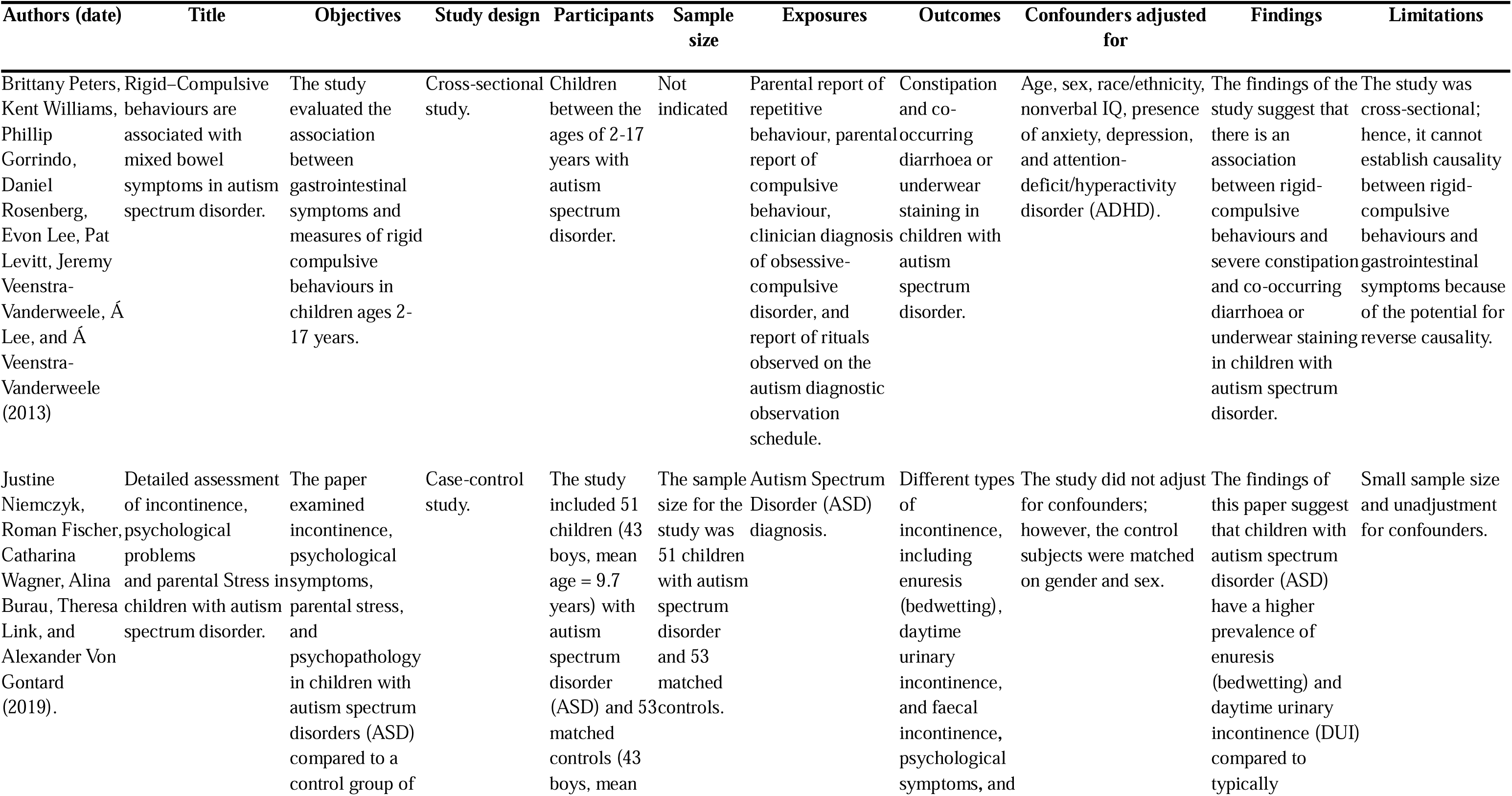
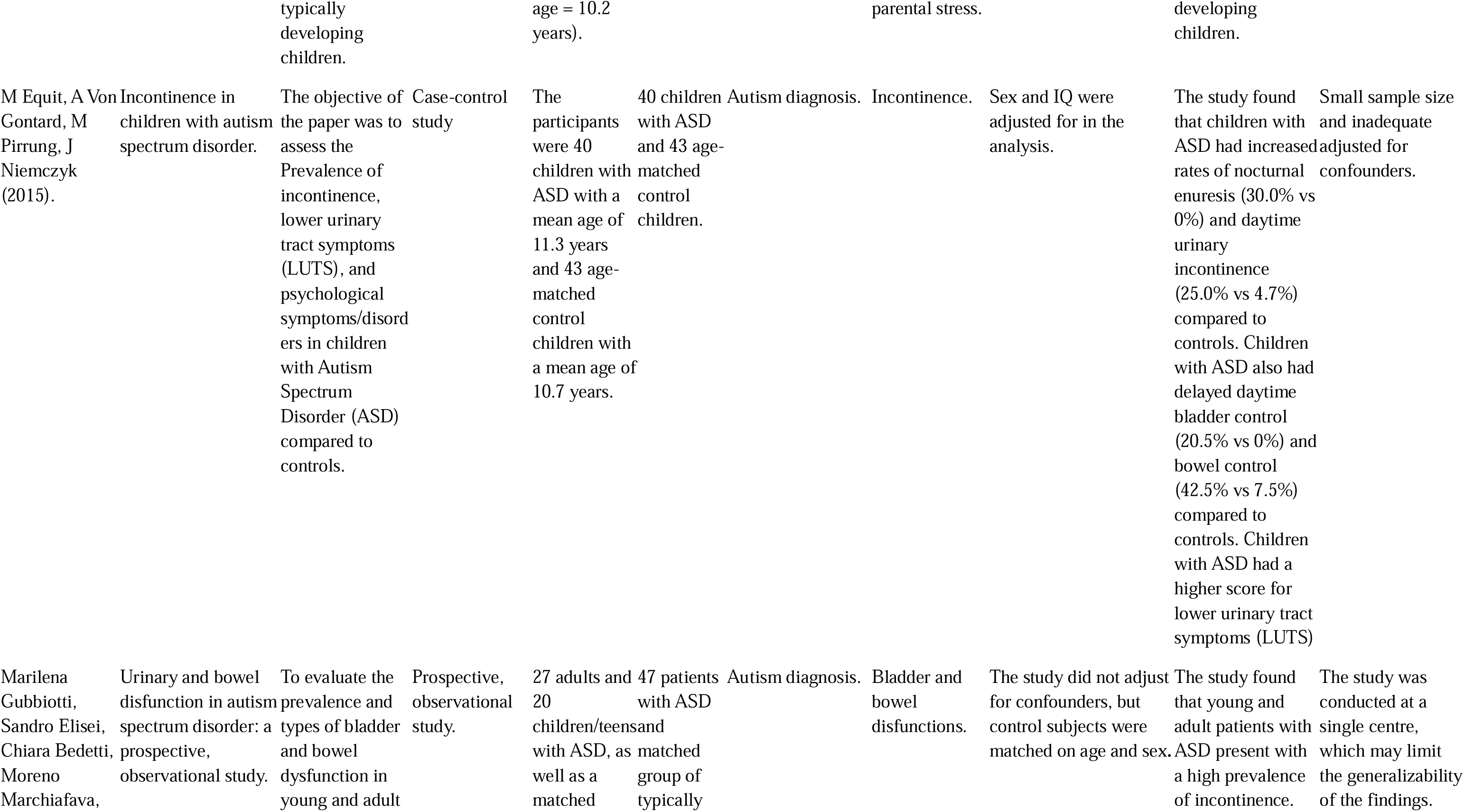
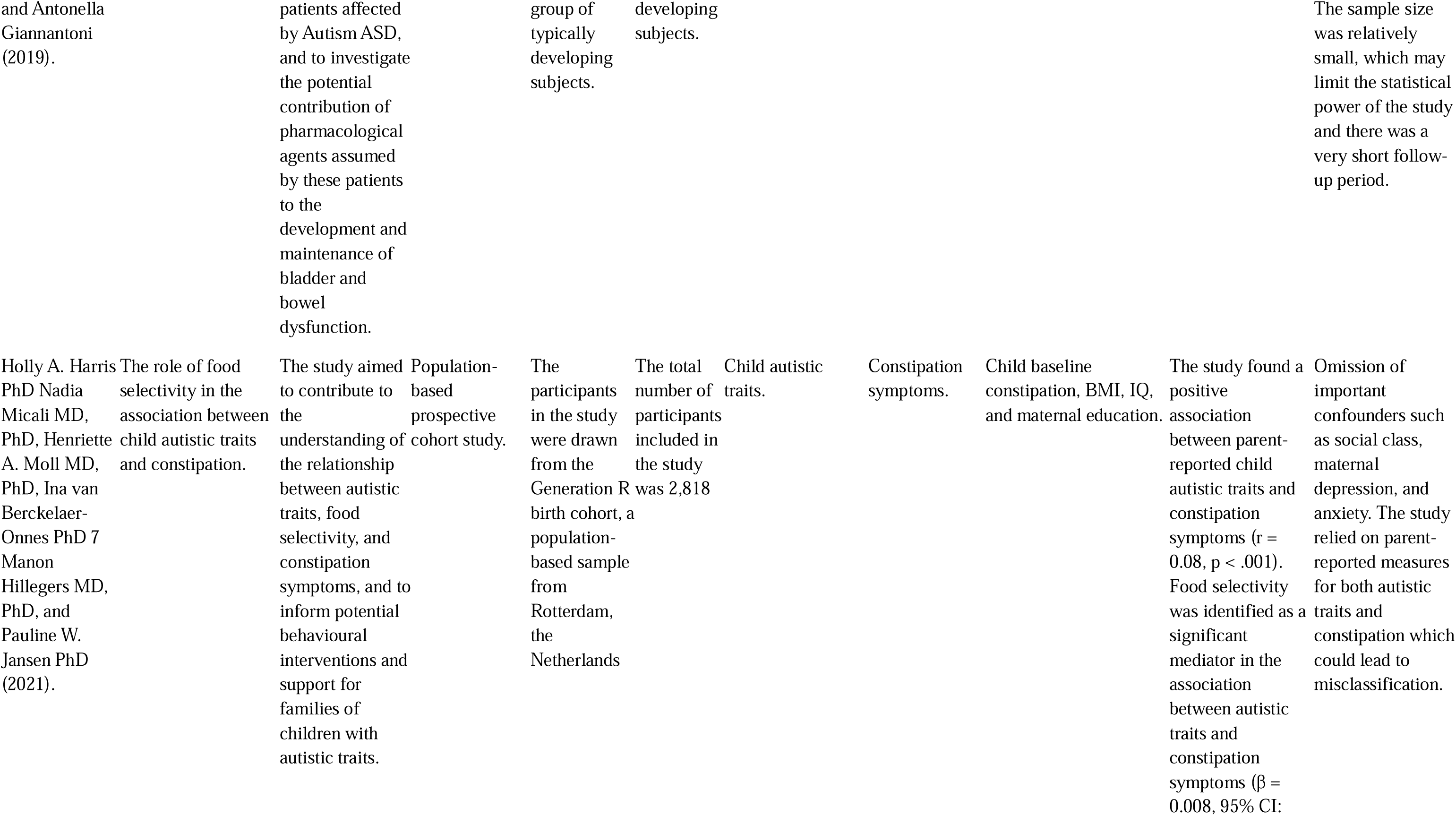
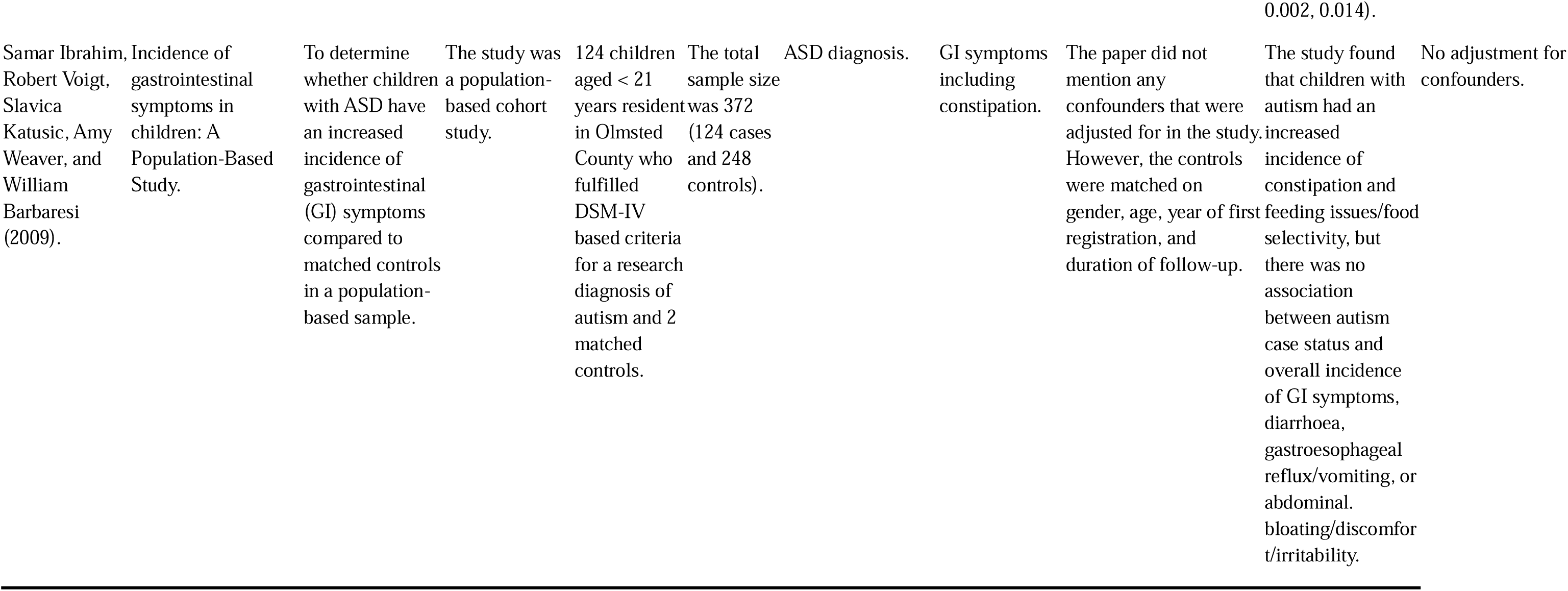
Characteristics of the studies included in the literature review.

**Supplementary table 2.** List of confounders, variable type, descriptions, and methods of assessment.

| Confounders | Variable type | Description | Method of assessment |
| --- | --- | --- | --- |
| Parity | Categorical | 0*<br>1<br>2 or more | Questionnaire |
| Child's sex | Binary | Female*<br>Male | Questionnaire |
| Family social class | Binary | Non-manual*<br>Manual | Questionnaire |
| Home ownership status | Binary | Mortgaged/owned*<br>Rented / other | Questionnaire |
| Mother highest educational attainment | Categorical | A-level or higher*<br>O-level<br>CSE/Vocational or less | Questionnaire |
| Maternal age at delivery | Continuous | 15-44 | Questionnaire |
| Child's developmental level at 18 months | Continuous score | -7.49 to 2.86 | Developed from the ALSPAC questionnaire |
| Maternal antenatal depression score (measured at 18 weeks and 32 weeks of gestation) | Continuous score | 0-30 | ALSPAC questionnaire using the Edinburgh Postnatal Depression Scale (EPDS) |
| Maternal postnatal depression score (measured at 8 weeks and 8 months after delivery) | Continuous score | 0-30 | ALSPAC questionnaire using the Edinburgh Postnatal Depression Scale (EPDS) |
| Maternal antenatal anxiety score (measured at 18 weeks of gestation) | Continuous score | 0-16 | ALSPAC questionnaire using the Crown-Crisp anxiety scale |
| Maternal postnatal anxiety score (measured at 8 weeks after delivery) | Continuous score | 0-16 | ALSPAC questionnaire using the Crown-Crisp anxiety scale |
| Financial difficulty score | Continuous score | 0-15 | ALSPAC questionnaire |

**Notes:**
ALSPAC-Avon Longitudinal Study of Parents and Children
\*Reference category

**Supplementary table 3.** The distribution of confounders and missing data across the samples shown in *Fig.1* for the age 9 years data.

| Variable | Initial number of participants in the study (<15645) | Participants with data for at least one autistic trait (15138) | Participants with data for at least one autistic trait and outcome variables (6708) | Participants with data for at least one autistic trait and outcome variables plus confounders (4233) |
| --- | --- | --- | --- | --- |
| **Confounders** | **N (%)** | **N (%)** | **N (%)** | **N (%)** |
| **SES** | **N = 11551** | **N = 11102** | **N = 6014** | **N = 4233** |
| Ref cat=non-manual |  |  |  |  |
| Manual | 2245 (19.44) | 2162(19.47) | 819 (13.62) | 528 (12.48) |
| Missing | 4094 (26.17) * | 4036 (26.66) * | 694 (10.35) * |  |
| **Home ownership** | **N = 9679** | **N = 9212** | **N = 5927** | **N = 4233** |
| Ref cat=Mortgaged/owned |  |  |  |  |
| Rented/Other | 1968 (20.33) | 1869 (20.29) | 902 (15.22) | 552 (13.05) |
| Missing | 5966 (38.13) * | 5926 (39.15) * | 781 (11.64) * |  |
| **Maternal education** | **N = 12470** | **N = 11982** | **N = 6277** | **N = 4233** |
| Ref cat=A-level or greater |  |  |  |  |
| O-level | 4319 (34.63) | 4156(34.69) | 2231 (35.54) | 1539 (36.37) |
| Certificate of Secondary Education, Vocational or less | 3753 (30.10) | 3600 (30.05) | 1302 (20.74) | 791 (18.70) |
| Missing | 3175 (20.29) * | 3156 (20.85) * | 431 (6.43) * |  |
| **Child sex** | **N = 15030** | **N = 14524** | **N =6708** | **N = 4233** |
| Ref cat=Female |  |  |  |  |
| Male | 7684 (51.12) | 7334 (50.49) | 3329 (49.63) | 2160 (51.05) |
| Missing | 615 (3.93) * | 614 (4.06) * |  |  |
| **Parity** | **N = 13101** | **N = 12613** | **N = 6252** | **N = 4233** |
| Ref cat=0 |  |  |  |  |
| 1 | 4576 (34.93) | 4391 (34.81) | 2231 (35.68) | 1502 (35.50) |
| 2 or more | 2661 (20.31) | 2553 (20.24) | 1059 (16.94) | 676 (15.98) |
| Missing | 2544 (16.26) * | 2525 (16.68) * | 456 (6.80) * |  |

**Notes:**
Ref cat-reference category
SES-Socioeconomic status
\*The proportions of missing values were calculated from the total number of participants for each column.

**Supplementary table 4.** The distribution of confounders and missing data across the samples shown in *Fig.1* for the age 14 years data.

| **Variable** | **Initial number of participants in the study (<15645)** | **Participants with data for at least one autistic trait (15138)** | **Participants with data for at least one autistic trait and outcome variables (5555)** | **Participants with data for at least one autistic trait and outcome variables plus confounders (3514)** |
| --- | --- | --- | --- | --- |
| **Confounders** | **N (%)** | **N (%)** | **N (%)** | **N (%)** |
| **SES** | **N = 11551** | **N = 11102** | **N = 4993** | **N = 3514** |
| Ref cat=non-manual |  |  |  |  |
| Manual | 2245 (19.44) | 2162(19.47) | 629 (12.60) | 410 (11.67) |
| Missing | 4094 (26.17) * | 4036 (26.66) * | 562 (10.12) * |  |
| **Home ownership** | **N = 9679** | **N = 9212** | **N = 4902** | **N = 3514** |
| Ref cat=Mortgaged/owned |  |  |  |  |
| Rented/other | 1968 (20.33) | 1869 (20.29) | 704 (14.36) | 441 (12.55) |
| Missing | 5966 (38.13) * | 5926 (39.15) * | 653 (11.76) * |  |
| **Maternal education** | **N = 12470** | **N = 11982** | **N = 5189** | **N = 3514** |
| Ref cat=A-level or greater |  |  |  |  |
| O-level | 4319 (34.63) | 4156(34.69) | 1819 (35.05) | 1261 (35.89) |
| Certificate of Secondary Education, Vocational or less | 3753 (30.10) | 3600 (30.05) | 988 (19.04) | 595 (16.93) |
| Missing | 3175 (20.29) * | 3156 (20.85) * | 366 (6.59) * |  |
| **Child sex** | **N = 15030** | **N = 14524** | **N = 5555** | **N = 3514** |
| Ref cat=Female |  |  |  |  |
| Male | 7684 (51.12) | 7334 (50.49) | 2570 (46.26) | 1695 (48.24) |
| Missing | 615 (3.93) * | 614 (4.06) * |  |  |
| **Parity** | **N = 13101** | **N = 12613** | **N = 5178** | **N = 3514** |
| Ref cat=0 |  |  |  |  |
| 1 | 4576 (34.93) | 4391 (34.81) | 1814 (35.03) | 1237 (35.20) |
| 2 or more | 2661 (20.31) | 2553 (20.24) | 857 (16.55) | 549 (15.62) |
| Missing | 2544 (16.26) * | 2525 (16.68) * | 377 (6.79) * |  |

**Notes:**
Ref cat-reference category
SES-Socioeconomic status
\*The proportions of missing values were calculated from the total number of participants for each column.

**Supplementary table 5.** The proportions of confounders, incontinence/constipation in children with each autistic trait/diagnosed autism for the age 9 years data.

| Variable | Social communications |  |  | Coherence |  |  | Sociability |  |  | Repetitive behaviour |  |  | Diagnosed autism |  |  | Factor-mean-score |  |  |
| --- | --- | --- | --- | --- | --- | --- | --- | --- | --- | --- | --- | --- | --- | --- | --- | --- | --- | --- |
|  | Non-cases<br>N (%) | Cases<br>N (%) | Chi<sup>2</sup><br>P-value | Non-cases<br>N (%) | cases<br>N (%) | Chi<sup>2</sup><br>P-value | Non-cases<br>N (%) | Cases<br>N (%) | Chi<sup>2</sup><br>P-value | Non-cases<br>N (%) | Cases<br>N (%) | Chi<sup>2</sup><br>P-value | Non-cases<br>N (%) | Cases<br>N (%) | Chi<sup>2</sup><br>P-value | Non-cases<br>N (%) | Cases<br>N (%) | Chi<sup>2</sup><br>P-value |
| **Confounders** |  |  |  |  |  |  |  |  |  |  |  |  |  |  |  |  |  |  |
| **Social class** |  |  |  |  |  |  |  |  |  |  |  |  |  |  |  |  |  |  |
| manual | 459<br>(11.97) | 63<br>(15.83) | 0.03 | 505<br>(12.44) | 62<br>(15.01) | 0.13 | 499<br>(12.76) | 55<br>(11.18) | 0.32 | 488<br>(12.34) | 31<br>(10.76) | 0.43 | 566<br>(12.77) | <5* | 0.09 | 510<br>(12.42) | 59<br>(15.32) | 0.10 |
| non-manual | 3,376<br>(88.03) | 335<br>(84.17) |  | 3,555<br>(87.56) | 351<br>(84.99) |  | 3,411<br>(87.24) | 437<br>(88.82) |  | 3,467<br>(87.66) | 257<br>(89.24) |  | 3,867<br>(87.23) | 54 |  | 3,595<br>(87.58) | 326<br>(84.68) |  |
| **Mum's highest educational level** |  |  |  |  |  |  |  |  |  |  |  |  |  |  |  |  |  |  |
| Certificate of Secondary Education, vocational or less | 703<br>(18.33) | 98<br>(24.62) | 0.01 | 757<br>(18.65) | 104<br>(25.18) | 0.004 | 727<br>(18.59) | 118<br>(23.98) | 0.003 | 736<br>(18.61) | 56<br>(19.44) | 0.92 | 858<br>(19.35) | 6<br>(10.53) | 0.24 | 775<br>(18.88) | 89<br>(23.12) | 0.10 |
| O-level | 1,387<br>(36.17) | 133<br>(33.42) |  | 1,476<br>(36.35) | 147<br>(35.59) |  | 1,415<br>(36.19) | 185<br>(37.60) |  | 1,440<br>(36.41) | 102<br>(35.42) |  | 1,607<br>(36.25) | 23<br>(40.35) |  | 1,491<br>(36.32) | 139<br>(36.10) |  |
| A-level or greater | 1,745<br>(45.50) | 167<br>(41.96) |  | 1,827<br>(45.00) | 162<br>(39.23) |  | 1,768<br>(38.41) | 189<br>(45.22) |  | 1,779<br>(44.98) | 130<br>(45.14) |  | 1,968<br>(44.39) | 28<br>(49.12) |  | 1,839<br>(44.80) | 157<br>(40.78) |  |
| **Home ownership** |  |  |  |  |  |  |  |  |  |  |  |  |  |  |  |  |  |  |
| rented/other | 464<br>(12.10) | 69<br>(17.34) | 0.003 | 521<br>(12.83) | 64<br>(15.50) | 0.13 | 506<br>(12.94) | 60<br>(12.20) | 0.641 | 484<br>(12.24) | 40<br>(13.89) | 0.41 | 588<br>(13.26) | <5* | 0.003 | 521<br>(12.69) | 67<br>(17.40) | 0.01 |
| mortgaged/owned | 3,371<br>(87.90) | 329<br>(82.66) |  | 3,539<br>(87.17) | 349<br>(84.50) |  | 3,404<br>(87.06) | 432<br>(87.80) |  | 3,471<br>(87.76) | 248<br>(86.11) |  | 3,845<br>(86.74) | 57 |  | 3,584<br>(87.31) | 318<br>(82.60) |  |
| **Parity** |  |  |  |  |  |  |  |  |  |  |  |  |  |  |  |  |  |  |
| 0 | 1,854<br>(48.34) | 186<br>(46.73) | 0.43 | 1,980<br>(48.77) | 171<br>(41.40) | 0.02 | 1,924<br>(49.21) | 194<br>(39.43) | <0.001 | 1,898<br>(47.99) | 139<br>(48.26) | 0.93 | 2,130<br>(48.05) | 27<br>(47.37) | 0.87 | 2,001<br>(48.75) | 156<br>(40.52) | 0.01 |
| 1 | 1,374<br>(35.83) | 139<br>(34.92) |  | 1,429<br>(35.20) | 170<br>(41.16) |  | 1,365<br>(34.91) | 208<br>(42.28) |  | 1,424<br>(36.01) | 101<br>(35.07) |  | 1,586<br>(35.78) | 22<br>(38.60) |  | 1,454<br>(35.42) | 154<br>(40.00) |  |
| 2 or more | 607<br>(15.83) | 73<br>(18.34) |  | 651<br>(16.03) | 72<br>(17.43) |  | 621<br>(15.88) | 90<br>(18.29) |  | 633<br>(16.01) | 48<br>(16.67) |  | 717<br>(16.17) | 8 (14.04) |  | 650<br>(15.83) | 75<br>(19.48) |  |
| **Child's sex** |  |  |  |  |  |  |  |  |  |  |  |  |  |  |  |  |  |  |
| female | 1,895<br>(49.41) | 138<br>(34.67) | <0.001 | 1,994<br>(49.11) | 146<br>(35.35) | <0.001 | 1,913<br>(48.93) | 197<br>(40.04) | <0.001 | 1,935<br>(48.93) | 106<br>(36.81) | <0.001 | 2,135<br>(48.16) | 14<br>(24.56) | <0.001 | 2,049<br>(49.91) | 100<br>(25.97) | <0.001 |
| male | 1940<br>(50.59) | 260<br>(65.33) |  | 2,066<br>(50.89) | 267<br>(64.65) |  | 1,997<br>(51.07) | 295<br>(59.96) |  | 2,020<br>(51.07) | 182<br>(63.19) |  | 2,298<br>(51.84) | 43<br>(75.44) |  | 2,056<br>(50.09) | 285<br>(74.03) |  |
| **Incontinence/constipation** |  |  |  |  |  |  |  |  |  |  |  |  |  |  |  |  |  |  |
| **Bedwetting** |  |  |  |  |  |  |  |  |  |  |  |  |  |  |  |  |  |  |
| Cases | 359<br>(9.36) | 62<br>(15.58) | <0.001 | 379<br>(9.33) | 61<br>(14.77) | <0.001 | 378<br>(9.67) | 57<br>(11.59) | 0.18 | 378<br>(9.56) | 37<br>(12.85) | 0.07 | 430<br>(9.70) | 11<br>(19.30) | 0.02 | 379<br>(9.23) | 62<br>(16.10) | <0.001 |
| Non-cases | 3,476<br>(90.64) | 336<br>(84.42) |  | 3,681<br>(90.67) | 352<br>(85.23) |  | 3,532<br>(90.33) | 435<br>(88.41) |  | 3,577<br>(90.44) | 251<br>(87.15) |  | 4,003<br>(90.30) | 46<br>(80.70) |  | 3,726<br>(90.77) | 323<br>(83.90) |  |
| **Daytime wetting** |  |  |  |  |  |  |  |  |  |  |  |  |  |  |  |  |  |  |
| Cases | 161<br>(4.20) | 39<br>(9.80) | <0.001 | 174<br>(4.29) | 40<br>(9.69) | <0.001 | 185<br>(4.73) | 24<br>(4.88) | 0.89 | 175<br>(4.42) | 20<br>(6.94) | 0.05 | 207<br>(4.67) | 8<br>(14.04) | 0.001 | 183<br>(4.46) | 32<br>(8.31) | 0.001 |
| Non-cases | 3,674<br>(95.80) | 359<br>(90.20) |  | 3,886<br>(95.71) | 373<br>(90.31) |  | 3,725<br>(95.27) | 468<br>(95.12) |  | 3,780<br>(95.58) | 268<br>(93.06) |  | 4,226<br>(95.33) | 49<br>(85.96) |  | 3,922<br>(95.54) | 353<br>(91.69) |  |
| **Soiling** |  |  |  |  |  |  |  |  |  |  |  |  |  |  |  |  |  |  |
| Cases | 177<br>(4.62) | 45<br>(11.31) | <0.001 | 186<br>(4.58) | 47<br>(11.38) | <0.001 | 196<br>(5.01) | 33<br>(6.71) | 0.11 | 196<br>(4.96) | 22<br>(7.64) | 0.05 | 228<br>(5.14) | 8<br>(14.04) | 0.003 | 185<br>(4.51) | 51<br>(13.25) | <0.001 |
| Non-cases | 3,658<br>(95.38) | 353<br>(88.69) |  | 3,874<br>(95.42) | 366<br>(88.62) |  | 3,714<br>(94.99) | 459<br>(93.29) |  | 3,759<br>(95.04) | 266<br>(92.36) |  | 4,205<br>(94.86) | 49<br>(85.96) |  | 3,920<br>(95.49) | 334<br>(86.75) |  |
| **Constipation** |  |  |  |  |  |  |  |  |  |  |  |  |  |  |  |  |  |  |
| Cases | 358<br>(9.34) | 47<br>(11.81) | 0.11 | 379<br>(9.33) | 44<br>(10.65) | 0.38 | 375<br>(9.59) | 41<br>(8.33) | 0.37 | 375<br>(9.48) | 29<br>(10.07) | 0.74 | 420<br>(9.47) | 5<br>(8.77) | 0.86 | 373<br>(9.09) | 52<br>(13.51) | 0.01 |
| Non-cases | 3,477<br>(90.66) | 351<br>(88.19) |  | 3,681<br>(90.67) | 369<br>(89.35) |  | 3,535<br>(90.41) | 451<br>(91.67) |  | 3,580<br>(90.52) | 259<br>(89.93) |  | 4,013<br>(90.53) | 52<br>(91.23) |  | 3,732<br>(90.91) | 333<br>(86.49) |  |

*Notes:* \*cell size may include zero or less than five, hence we did not present the percentages.
For social communication and coherence, cases were those in the worst 10% of the SCDC score and the coherence subscale respectively. For sociability, cases were those in the worst 11% score of the sociability scale. Cases for repetitive behaviours were those in the worst 7% of the repetitive behaviour scale. Non-cases were individuals below the thresholds indicated for each trait. For diagnosed autism, cases were those who had a confirmed diagnosis of autism while non-cases were those who did not have a confirmed diagnosis. For the factor-mean-score, cases were those in the worst 10% score.

**Supplementary table 6.** The proportions of confounders, incontinence/constipation in children with each autistic trait/diagnosed autism for the age 14 years data.

| Variable | Social-communication difficulties |  |  | Coherence difficulties |  |  | Sociability difficulties |  |  | Repetitive behaviour |  |  | Diagnosed autism |  |  | Factor-mean-scores |  |  |
| --- | --- | --- | --- | --- | --- | --- | --- | --- | --- | --- | --- | --- | --- | --- | --- | --- | --- | --- |
|  | Non-cases<br>N (%) | Cases<br>N (%) | Chi<sup>2</sup><br>P-value | Non-cases<br>N (%) | cases<br>N (%) | Chi<sup>2</sup><br>P-value | Non-cases<br>N (%) | Cases<br>N (%) | Chi<sup>2</sup><br>P-value | Non-cases<br>N (%) | Cases<br>N (%) | Chi<sup>2</sup><br>P-value | Non-cases<br>N (%) | Cases<br>N (%) | Chi<sup>2</sup><br>P-value | Non-cases<br>N (%) | Cases<br>N (%) | Chi<sup>2</sup><br>P-value |
| **Confounders** |  |  |  |  |  |  |  |  |  |  |  |  |  |  |  |  |  |  |
| **Social class** |  |  |  |  |  |  |  |  |  |  |  |  |  |  |  |  |  |  |
| manual | 344<br>(10.84) | 39<br>(14.03) | 0.11 | 348<br>(11.01) | 38<br>(12.42) | 0.46 | 380<br>(11.74) | 37<br>(9.81) | 0.27 | 370<br>(11.32) | 24<br>(11.06) | 0.91 | 432<br>(11.79) | <5* | 0.13 | 384<br>(11.25) | 49<br>(17.25) | 0.003 |
| non-manual | 2,829<br>(89.16) | 239<br>(85.97) |  | 2,812<br>(88.99) | 268<br>(87.58) |  | 2,857<br>(88.26) | 340<br>(90.19) |  | 2,899<br>(88.68) | 193<br>(88.94) |  | 3,233<br>(88.21) | 31 |  | 3,029<br>88.75 | 235<br>82.75 |  |
| **Mum's highest educational level** |  |  |  |  |  |  |  |  |  |  |  |  |  |  |  |  |  |  |
| Certificate of Secondary Education, vocational or less | 512<br>(16.14) | 55<br>(19.78) | 0.20 | 512<br>(16.20) | 68<br>(22.22) | 0.03 | 543<br>(16.77) | 76<br>(20.16) | 0.14 | 544<br>(16.64) | 38<br>(17.51) | 0.93 | 630<br>(17.19) | 6<br>(18.75) | 0.24 | 570<br>(16.70) | 66<br>(23.24) | 0.02 |
| O-level | 1,141<br>(35.96) | 89<br>(32.01) |  | 1,123<br>(35.54) | 104<br>(33.99) |  | 1,153<br>(35.62) | 139<br>(36.87) |  | 1,175<br>(35.94) | 76<br>(35.02) |  | 1,314<br>(35.85) | 7<br>(21.88) |  | 1,231<br>(36.07) | 90<br>(31.69) |  |
| A-level or greater | 1,520<br>(47.90) | 134<br>(48.20) |  | 1,525<br>(48.26) | 134<br>(43.79) |  | 1,541<br>(47.61) | 162<br>(42.97) |  | 1,550<br>(47.42) | 103<br>(47.47) |  | 1,721<br>(46.96) | 19<br>(59.38) |  | 1,612<br>(47.23) | 128<br>(45.07) |  |
| **Home status** |  |  |  |  |  |  |  |  |  |  |  |  |  |  |  |  |  |  |
| rented/other | 353<br>(11.13) | 42<br>(15.11) | 0.05 | 361<br>(11.42) | 38<br>(12.42) | 0.60 | 399<br>(12.33) | 36<br>(9.55) | 0.12 | 379<br>(11.59) | 34<br>(15.67) | 0.07 | 459<br>(12.52) | <5* | 0.03 | 420<br>(12.31) | 39<br>(13.73) | 0.48 |
| mortgaged/owned | 2,820<br>(88.87) | 236<br>(84.89) |  | 2,799<br>(88.58) | 268<br>(87.58) |  | 2,838<br>87.67 | 341<br>90.45 |  | 2,890<br>(88.41) | 183<br>(84.33) |  | 3,206<br>(87.48) | 32 |  | 2,993<br>(87.69) | 245<br>(86.27) |  |
| **Parity** |  |  |  |  |  |  |  |  |  |  |  |  |  |  |  |  |  |  |
| 0 | 1,562<br>(49.23) | 133<br>(47.84) | 0.83 | 1,563<br>(49.46) | 130<br>(42.48) | 0.04 | 1,620<br>(50.05) | 154<br>(40.85) | 0.002 | 1,592<br>(48.70) | 112<br>(51.61) | 0.60 | 1,788<br>(48.79) | 18 | 0.56 | 1,685<br>(49.37) | 121<br>(42.61) | 0.07 |
| 1 | 1,126<br>(35.49) | 99<br>(35.61) |  | 1,102<br>(34.87) | 128<br>(41.83) |  | 1,114<br>(34.41) | 162<br>(42.97) |  | 1,169<br>(35.76) | 76<br>(35.02) |  | 1,304<br>(35.58) | 11 |  | 1,205<br>(35.31) | 110<br>(38.73) |  |
| 2 or more | 485<br>(15.29) | 46<br>(16.55) |  | 495<br>(15.66) | 48<br>(15.69) |  | 503<br>(15.54) | 61<br>(16.18) |  | 508<br>(15.54) | 29<br>(13.36) |  | 573<br>(15.63) | <5* |  | 1,685<br>(49.37) | 121<br>(42.61) |  |
| **Child's sex** |  |  |  |  |  |  |  |  |  |  |  |  |  |  |  |  |  |  |
| female | 1,648 | 109 | <0.001 | 1,648 | 113 | <0.001 | 1,668 | 167 | 0.01 | 1,686 | 87 | 0.001 | 1,870 | 9 | 0.01 | 1,803 | 76 | <0.001 |
|  | (51.94) | (39.21) |  | (52.15) | (36.93) |  | (51.53) | (44.30) |  | 51.58 | 40.09 |  | (51.02) | (28.13) |  | (52.83) | (26.76) |  |
| male | 1,525 | 169 |  | 1,512 | 193 |  | 1,569 | 210 |  | 1,583 | 130 |  | 1,795 | 23 |  | 1,610 | 208 |  |
|  | (48.06) | (60.79) |  | (47.85) | (63.07) |  | 48.47 | 55.70 |  | 48.42 | 59.91 |  | (48.98) | (71.88) |  | (47.17) | (73.24) |  |
| **Incontinence/constipation** |  |  |  |  |  |  |  |  |  |  |  |  |  |  |  |  |  |  |
| **Bedwetting** |  |  |  |  |  |  |  |  |  |  |  |  |  |  |  |  |  |  |
| Cases | 74 | 8 | 0.57 | 69 | 11 | 0.12 | 75 | 9 | 0.93 | 80 | <5* | 0.15 | 85 | <5* | 0.76 | 70 | 16 | <0.001 |
|  | (2.33) | (2.88) |  | (2.18) | (3.59) |  | (2.32) | (2.39) |  | (2.45) |  |  | (2.32) |  |  | (2.05) | (5.63) |  |
| Non-cases | 3,099 | 270 |  | 3,091 | 295 |  | 3,162 | 368 |  | 3,189 | 215 |  | 3,580 | 31 |  | 3,343 | 268 |  |
|  | (97.67) | (97.12) |  | (97.82) | (96.41) |  | (97.68) | (97.61) |  | (97.55) |  |  | (97.68) |  |  | (97.95) | (94.37) |  |
| **Daytime wetting** |  |  |  |  |  |  |  |  |  |  |  |  |  |  |  |  |  |  |
| Cases | 88 | 9 | 0.65 | 89 | 12 | 0.27 | 93 | 11 | 0.96 | 94 | 5 | 0.62 | 106 | <5* | 0.94 | 92 | 15 | 0.01 |
|  | (2.77) | (3.24) |  | (2.82) | (3.92) |  | (2.87) | (2.92) |  | (2.88) | (2.30) |  | (2.89) |  |  | (2.70) | (5.28) |  |
| Non-cases | 3,085 | 269 |  | 3,071 | 294 |  | 3,144 | 366 |  | 3,175 | 212 |  | 3,559 | 31 |  | 3,321 | 269 |  |
|  | (97.23) | (96.76) |  | (97.18) | (96.08) |  | (97.13) | (97.08) |  | (97.12) | (97.70) |  | (97.11) |  |  | (97.30) | (94.72) |  |
| **Soiling** |  |  |  |  |  |  |  |  |  |  |  |  |  |  |  |  |  |  |
| Cases | 137 | 18 | 0.10 | 133 | 24 | 0.004 | 137 | 24 | 0.06 | 145 | 13 | 0.29 | 161 | 5 | 0.02 | 138 | 28 | <0.001 |
|  | (4.32) | (6.47) |  | (4.21) | (7.84) |  | (4.23) | (6.37) |  | (4.44) | (5.99) |  | (4.39) | (15.63) |  | (4.04) | (9.86) |  |
| Non-cases | 3,036 | 260 |  | 3,027 | 282 |  | 3,100 | 353 |  | 3,124 | 204 |  | 3,504 | 27 |  | 3,275 | 256 |  |
|  | (95.68) | (93.53) |  | (95.79) | (92.16) |  | (95.77) | (93.63) |  | (95.56) | (94.01) |  | (95.61) | (84.38) |  | (95.96) | (90.14) |  |
| **Constipation** |  |  |  |  |  |  |  |  |  |  |  |  |  |  |  |  |  |  |
| Cases | 216 | 34 | 0.001 | 217 | 37 | 0.001 | 227 | 35 | 0.11 | 227 | 30 | 0.001 | 265 | 5 | 0.07 | 236 | 34 | 0.002 |
|  | (6.81) | (12.23) |  | (6.87) | (12.09) |  | (7.01) | (9.28) |  | (6.94) | (13.82) |  | (7.23) | (15.63) |  | (6.91) | (11.97) |  |
| Non-cases | 2,957 | 244 |  | 2,943 | 269 |  | 3,010 | 342 |  | 3,042 | 187 |  | 3,400 | 27 |  | 3,177 | 250 |  |
|  | 93.19 | 87.77 |  | (93.13) | (87.91) |  | (92.99) | (90.72) |  | (93.06) | (86.18) |  | (92.77) | (84.38) |  | (93.09) | (88.03) |  |

Notes: \*cell size may include zero or less than five, hence we did not present the percentages.
For social communication and coherence, cases were those in the worst 10% of the SCDC score and the coherence subscale respectively. For sociability, cases were those in the worst 11% score of the sociability scale. Cases for repetitive behaviours were those in the worst 7% of the repetitive behaviour scale. Non-cases were individuals below the thresholds indicated for each trait. For diagnosed autism, cases were those who had a confirmed diagnosis of autism while non-cases were those who did not have a confirmed diagnosis. For the factor-mean-score, cases were those in the worst 10% score.

